# Creating and validating a DNA methylation-based proxy for Interleukin-6

**DOI:** 10.1101/2020.07.20.20156935

**Authors:** Anna J. Stevenson, Danni A. Gadd, Robert F. Hillary, Daniel L. McCartney, Archie Campbell, Rosie M. Walker, Kathryn L. Evans, Sarah E. Harris, Tara L Spires-Jones, Allan F. MacRae, Peter M. Visscher, Andrew M McIntosh, Ian J Deary, Riccardo E Marioni

## Abstract

Chronic inflammation is a pervasive feature of ageing and may be linked to age-related cognitive decline. However, population studies evaluating its relationship with cognitive functioning have produced heterogeneous results. A potential reason for this is the variability of inflammatory mediators which could lead to misclassifications of individuals’ persisting levels of inflammation. The epigenetic mechanism DNA methylation has shown utility in indexing environmental exposures and could potentially be leveraged to provide proxy signatures of chronic inflammation.

We conducted an elastic net regression of interleukin-6 (IL-6) in a cohort of 895 older adults (mean age: 69 years) to develop a DNA methylation-based predictor. The predictor was tested in an independent cohort (n=7,028 [417 with measured IL-6], mean age: 51 years).We examined the association between the DNA methylation IL-6 score and serum IL-6, its association with age and established correlates of circulating IL-6, and with cognitive ability.

A weighted score from 12 DNA methylation sites optimally predicted IL-6 (independent test set R^2^=5.1%). In the independent test cohort, both measured IL-6, and the DNA methylation proxy, increased as a function of age (serum IL-6: n=417, β=0.02, SE=0.004 p=1.3×10^−7^; DNAm IL-6 score: n=7,028, β=0.02, SE=0.0009, p<2 × 10^−16^). Serum IL-6 was not found to associate with cognitive ability (n=417, β=-0.06, SE=0.05, p=0.19); however, an inverse association was identified between the DNA methylation score and cognitive functioning (n=7,028, β=-0.14, SE=0.02, p_FDR_=1.5 × 10^−14^).

These results suggest DNA methylation-based predictors can be used as proxies for inflammatory markers, potentially allowing for reliable insights into the relationship between chronic inflammation and pertinent health outcomes.

## 1. INTRODUCTION

Acute inflammation is a necessary component of the biological response to harmful stimuli. However, there is increasing recognition that a chronic, sub-acute elevation of serum concentrations of pro-inflammatory mediators occurring in older age may underpin the development of many age-related diseases, including cancer, atherosclerosis and Alzheimer’s disease (1-4). In spite of this, studies of chronic inflammation and its therapeutics, particularly in cognitive decline and dementia, have produced eminently heterogeneous results (5, 6).

Population studies investigating the relationship between chronic inflammation and incident morbidities typically rely on a single measurement of circulating inflammatory biomarker levels as proxies for individuals’ persistent inflammatory states. Interleukin-6 (IL-6), a pleiotropic pro-inflammatory cytokine, is a principal stimulator of an extensive range of acute-phase inflammatory proteins, including C-reactive protein (CRP), serum amyloid A and fibrinogen (7). It has been hypothesised that IL-6 is critical in the transition from the acute, beneficial inflammatory response, to chronic and deleterious inflammation, marking it as a key target for research into the process (8). However, inflammatory cytokines are vulnerable to analytical measurement error; specifically, circulating plasma IL-6 levels can show transient variability due to manifold influences, including the obvious precipitate – infection - but also diet and activity levels (9, 10). This is an important consideration when utilising it as a surrogate of chronic inflammation (11, 12).

Epigenetic mechanisms afford an opportunity to further understand the molecular pathophysiology of inflammation. DNA methylation (DNAm), the most commonly-studied epigenetic mechanism, is involved in the regulation of gene expression and chromosomal stability (13). It has been proposed as a putative intermediary, linking inflammation with sequent disease outcomes, with differential DNAm profiles identified in inflammatory diseases (14-16). Studies are beginning to focus on resolving the link between inflammation and DNAm (17-19); however, there remains a relative dearth of understanding about the association between DNAm and pro-inflammatory cytokines, and of how this information could be used to inform disease progression or risk stratification. Recently, DNAm alterations have been leveraged to provide peripheral predictors, or biomarkers, of disease status and as archives of habitual traits. These indices have demonstrated potential utility in epidemiological research contexts (20).

In this study, we utilise DNAm data from the Lothian Birth Cohort 1936 to train an optimised predictor of IL-6 using a penalised regression model. This DNAm IL-6 score was subsequently applied to a large out-of-sample cohort (Generation Scotland) to (i) characterise its relationship with measured IL-6 and IL-6 correlates; and (ii) determine its association with cognitive ability, a trait correlated with a variety of health outcomes.

## 2. METHODS

### 2.1 The Lothian Birth Cohort 1936

The Lothian Birth Cohort 1936 (LBC1936) is a longitudinal study of ageing comprising individuals born in 1936. Most of them took part in the Scottish Mental Survey 1947 in which a test of cognitive ability was administered to children attending school in Scotland at age around 11 years.

Participants living in Edinburgh and the Lothians were re-contacted around 60 years later, with 1,091 individuals consenting to join the LBC1936 study. At recruitment, participants were aged around 70 years and subsequently have completed up to four waves of detailed testing, repeated triennially. Comprehensive genetic, epigenetic, cognitive, psycho-social, health and lifestyle, and biomarker data are available for these individuals across the eighth decade. Full details of the recruitment and testing protocol for the study has been described previously (21, 22). Ethical permission for the LBC1936 was obtained from the Multi-Centre Research Ethics Committee for Scotland (MREC/01/0/56) and the Lothian Research Ethics Committee (LREC/2003/2/29). Written informed consent was obtained from all participants.

#### 2.1.1 LBC1936 DNA methylation preparation

DNA extracted from whole blood was analysed using the Illumina 450K BeadChip methylation array at the Edinburgh Clinical Research Facility. Details of quality control procedures have been described previously (23, 24). Briefly, raw intensity data was background-corrected and normalised using internal controls. Low-quality samples (those exhibiting inadequate bisulphite conversion, hybridisation or nucleotide extension) were removed upon manual inspection. Quality control further removed probes with a low detection rate (P>0.01) and samples with a low call rate (<450,000 probes detected at p-value <0.01). Finally, samples with a mismatch between genotype and SNP control probes or DNAm-predicted and reported sex, were additionally removed.

Methylation data was subset to probes in common on both the 450k and EPIC arrays (n=428,489). DNAm data was available for 889 individuals at Wave 1 of the study (aged ∼70 years).

#### 2.1.2 Phenotype preparation

Blood plasma samples were collected from LBC1936 participants at Wave 1 of the study and were analysed using a 92-plex proximity extension assay (Olink^®^ Bioscience, Uppsala, Sweden). This represents the Olink^®^ inflammation panel which provides a multiplex immunoassay, facilitating the analysis of 92 inflammation-related protein biomarkers. We focused analyses on IL-6 as this was measured in the prediction cohort (see section 4.2) and is widely used in studies of inflammation and cognitive function. Briefly, the Olink^®^ protocol involves the incubation of 1μl of plasma sample with proximity antibody pairs linked to DNA reporter molecules. Once a complementary antibody has bound to the antibody pair, the DNA tails form an amplicon by proximity extension which is subsequently quantified using high-throughput real-time polymerase chain reaction. Data underwent pre-processing by Olink^®^ using NPX Manager software. Prior to analyses, IL-6 levels were rank-based inverse normalised to correct for skewness, and normalised protein levels were then regressed onto age, sex, four genetic principal components and Olink^®^ array plate. Standardised residuals from this regression model were used in the penalised regression analysis. Complete DNAm and IL-6 data was available for 875 individuals.

#### 2.1.3 Development of a DNA methylation based predictor of IL-6 in LBC1936

An elastic net penalised regression was run to derive the DNA methylation-based predictor of IL-6. This method helps to control for collinearity in predictor variables and is particularly useful when dealing with a large number of predictors. The elastic net penalty is an intermediate of LASSO and ridge regression - the LASSO approach yields a sparse solution with a minimal set of non-zero coefficients from the feature set (it will include one feature from a set of correlated features) and ridge regression shrinks the coefficients for correlated features towards each other (so includes a large number of features).

The elastic net model was run using the *glmnet* library in R (25), with twelve-fold cross-validation. To reduce overfitting, folds were specified by methylation analysis batch, yielding between 63 and 83 observations per fold. IL-6 and DNA methylation levels at 428,489 CpG sites were entered as the dependent and independent variables, respectively. In order to apply the relevant penalty, the mixing parameter (α) was set to 0.5. Coefficients for the model with the lambda value (regularisation parameter) corresponding to the minimum mean cross-validated error were extracted. These weights were then applied to the corresponding CpGs in an out-of-sample prediction cohort in order to derive the DNAm-based predictor of IL-6.

### 2.2 Out-of-sample prediction cohort: Generation Scotland

The Generation Scotland (GS) cohort was used for external predictions. GS is a family-structured, population-based genetic epidemiology cohort comprising ∼22,000 participants aged between 18 and 99 years. These individuals have been widely profiled in regards to cognitive ability, health, genetics and epigenetics, further details of which can be found elsewhere (26). All components of GS received ethical approval from the NHS Tayside Committee on Medical Research Ethics (REC Reference Number: 05/S1401/89). GS has also been granted Research Tissue Bank status by the Tayside Committee on Medical Research Ethics (REC Reference Number: 10/S1402/20), providing generic ethical approval for a wide range of uses within medical research.

#### 2.2.1 DNA methylation in GS

The DNA methylation arrays in GS were run in two separate sets. The present study includes analysis of methylation data from 7,028 unrelated individuals in total (2,578 in the first set and 4,450 in the second set). Methylation was quantified from whole-blood using the Illumina HumanMethylationEPIC BeadChip. Quality control steps for set 1 have been reported previously (27). Briefly, samples were removed (i) if they were deemed as outliers based on a visual inspection of the log median intensity of the methylated versus un-methylated signal per array, (ii) if there was a mismatch between their predicted- and reported sex and/or (iii) if ≥1% of CpGs had a detection p-value >0.05. Probes were removed based on (ii) a bead count <3 in >5% of samples, and (ii) ≥5% of samples having a detection p-value >0.05. CpGs with missing values, non-CpG probes and non-autosomal probes were additionally removed. Quality control steps for set 2 have also been reported previously and were near identical to the set 1 protocol (28).

#### 2.2.2 Phenotype preparation in GS

##### Serum IL-6

Serum IL-6 was quantified at the University of Glasgow using a high-sensitivity commercial enzyme-linked immunosorbent assay (R&D systems, Oxon, UK). IL-6 data was available for 417 individuals. These samples had previously been selected as father/offspring pairs in a telomere length study. The data in the current study was subset to exclude any related individuals. This limited the number of samples from older females in the dataset. To correct for positive skew, IL-6 data was log-transformed (natural log) prior to analyses.

##### Correlates of IL-6

Several modifiable and non-modifiable factors have been shown to associate with circulating IL-6 levels, including smoking (29, 30), alcohol intake (31), body mass index (BMI; (32, 33)) and socioeconomic status (34, 35). These were assessed in GS as follows: smoking was divided into current smokers, ex-smokers (split into those who had quit within the previous 12 months of their blood sample date, and those who had quit prior to that); alcohol consumption (units) in a typical week; BMI was calculated as the ratio of weight in kg divided by height in m^2^; and social deprivation was measured using the Scottish Index of Multiple Deprivation (SIMD - www.simd.scot). The SIMD ranks geographical areas in Scotland based on current income, employment, health, education, skills and training, geographic access to services, housing and crime. The SIMD provides a standardised measure of relative deprivation throughout Scotland. To reduce positive skew, a log(units+1) transformation was used for the alcohol consumption data prior to analysis.

Due to the inclusion of CpG sites that have previously been related to smoking in the DNAm IL-6 score, we additionally generated a methylation-based smoking score using the *EpiSmokEr* R package (36). *EpiSmokEr* provides a smoking score derived from methylation levels at 187 CpG sites based on an EWAS of current v. never smokers (37, 38).

##### Cognitive tests

To derive a general cognitive ability phenotype, a principal component analysis was applied to the cognitive tests of executive function (verbal fluency), processing speed (digit-symbol coding) and logical memory. Scores on the first un-rotated principal component were used as a general fluid-type cognitive ability score for each individual in GS. This component accounted for 50% of the variance and individual test loadings ranged from 0.63 to 0.77. Full details on the cognitive assessments for the GS cohort have been described previously (39, 40).

### 2.3 Statistical analysis

Pearson correlations were calculated between serum IL-6 and the DNAm IL-6 score and between the latter and the imputed immune cell proportions (CD8+T cells, CD4+T cells, Natural Killer cells, B cells, Monocytes and Granulocytes). The relationship between IL-6 and the estimated immune cell proportions was further investigated in linear regression models. Here, the difference between the R^2^ statistic from a basic model (IL-6 ∼ age + sex), and the same model adjusted for the imputed immune cell proportions, was calculated to estimate the proportion of variance accounted for by these predictors.

To investigate the validity of the DNAm IL-6 score, linear regression models were run with phenotypes that have previously been associated with circulating IL-6 levels: BMI, smoking, alcohol intake, and social deprivation. Measured IL-6 models were adjusted for age, sex and methylation set with the DNAm IL-6 models further adjusted for cell proportions imputed from the methylation data. In a sensitivity analysis, the models were re-run adjusting for methylation-predicted smoking score using the *EpiSmokEr* output.

Linear regression models were used to establish the association with age of both measured IL-6 and the DNAm IL-6 score, and their associations with cognitive ability. Sex, methylation set and estimated cell proportions were included as covariates in both models. The cognitive ability model was further adjusted for age and, as above, was repeated adjusting for methylation smoking score to test for confounding. All analyses were conducted in the R statistical environment version 3.5.0 (41). Correction for multiple testing was applied using the false discovery rate (FDR p<0.05).

## 3. RESULTS

### 3.1 Cohort information

Summary information for all variables included in analyses is presented in **Table 1**. LBC1936 is an older cohort than GS (LBC1936 Wave 1 mean age: 69.6 years; GS mean age: 50.8 years) with a more even balance of the sexes (LBC1936: 49.4% female; GS: 58.2% female).

**Table 1.** Summary of the LBC1936 (Wave 1) and GS variables.

|  | LBC1936 |  |  | GS |  |  |
| --- | --- | --- | --- | --- | --- | --- |
|  | n | mean | SD | n | mean | SD |
| *Age (years)* | 895 | 69.6 | 0.8 | 7,028 | 50.8 | 12.9 |
| *Females (%)* | 437 (49.9) | - | - | 4,089 (58.2) | - | - |
| *IL-6 (pg/mL)* | 875 | 3.99 | 1.87 | 417 | 1.8 | 1.9 |
| *Alcohol (average units per week)* | - | - | - | 6,414 | 10.5 | 11.02 |
| *BMI (kg/m<sup>2</sup>)* | - | - | - | 6,988 | 26.9 | 5.09 |
| *Smoking status (%)* | - | - | - | 6,854 | - | - |
| *-Current* |  |  |  | 1,135 (16%) | - | - |
| *-Ex* |  |  |  | 2,167 (32%) | - | - |
| *-Never* |  |  |  | 3,552 (52%) | - | - |
| *SIMD* | - | - | - | 6,673 | 3953 | 1858 |

BMI=body mass index; SIMD=Scottish Index of Multiple Deprivation

### 3.2 Elastic net regression in LBC1936

The elastic net model returned 12 CpG sites. The probes and their relevant weights are presented in **Supplementary Table 1**. A DNAm proxy for IL-6 was generated in GS by multiplying methylation beta values with the coefficients from this output, and summing into a single score.

### 3.3 Correlation with IL-6 in GS

To test the predictive performance, Pearson correlations were calculated between the DNAm IL-6 score and measured IL-6 in GS. The correlation coefficient between measured log(IL-6) and the elastic net-derived IL-6 score was 0.23 (variance explained = 5.1%). A plot of serum IL-6 versus this DNAm IL-6 score is presented in **Figure 1**.

**Figure 1.**
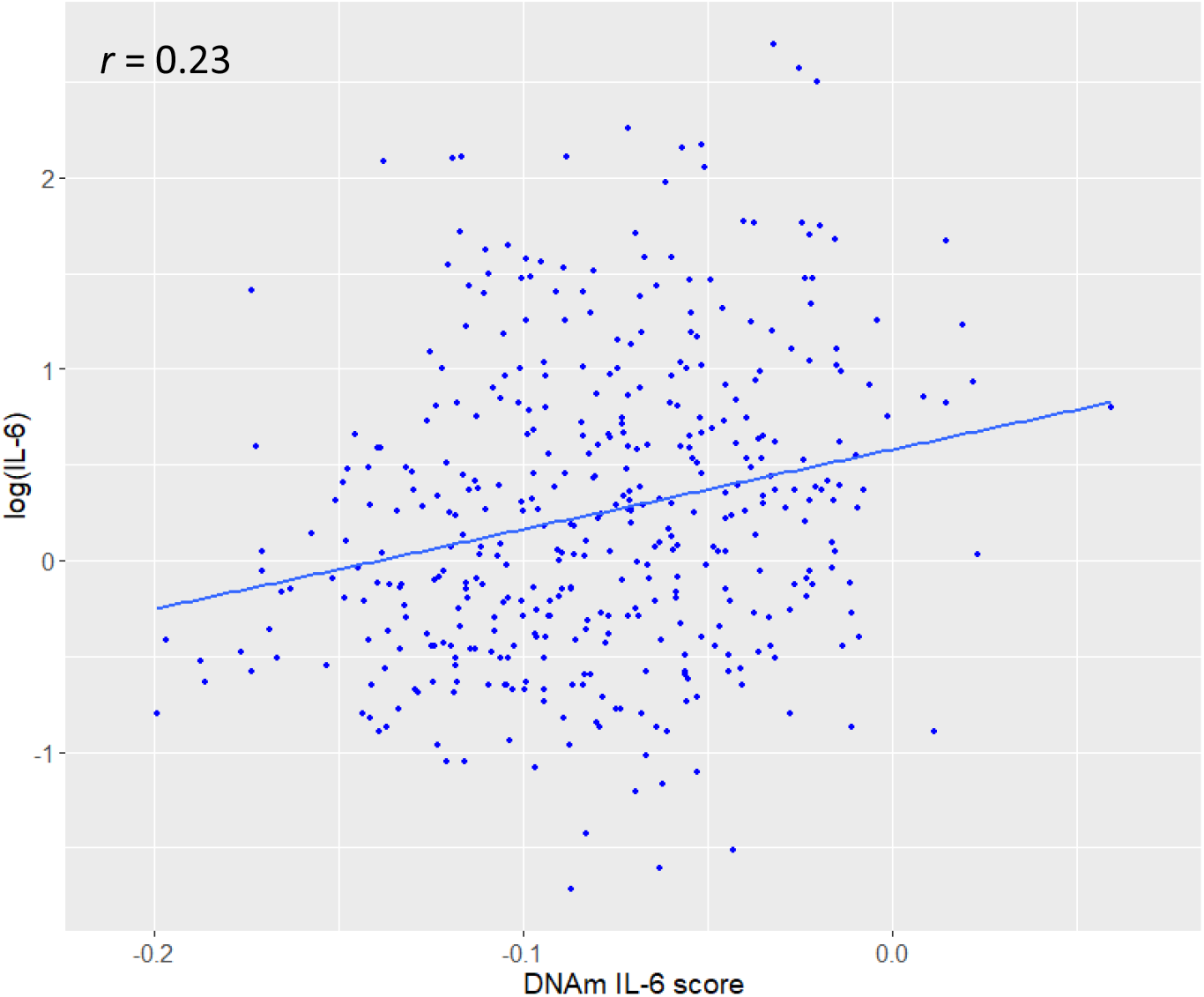
Plot of the relationship between log(IL-6) and the DNAm IL-6 score in the subset of the Generation Scotland dataset in which serum IL-6 was measured (n=417).

Methylation at one of the probes included in the score (cg05575921, *AHRR*), has consistently been strongly inversely associated with smoking status (42-45), with four further probes (cg25250132, cg17412005, cg25250132, cg20059928, cg04928129) showing similar, though weaker, associations (46, 47). Because of this, the association between the DNAm IL-6 score and log(IL-6) was re-run in a regression analysis, adjusting for a methylation-based score for smoking. In the unadjusted model the DNAm IL-6 score was significantly positively associated with log(IL-6) (β=0.18, SE=0.04, p=2.9 × 10^− 6^). Adjusting for the methylation smoking score had little effect on this relationship (β=0.17, SE=0.04, p=1.4 × 10^−5^), suggesting the association between measured IL-6 and the DNAm IL-6 score is largely not influenced by smoking. A correlation plot of log(IL-6), the DNAm IL-6 score and the methylation-predicted smoking score is presented in **Supplementary Figure 2**.

### 3.4 Correlation with cell proportions in LBC1936 and GS

The correlation between the DNAm IL-6 score and the estimated leukocyte proportions is presented in **Supplementary Figure 1**. These ranged from −0.69 (CD4^+^T cells) to 0.49 (Granulocytes). Due to the high correlations identified here, we additionally tested the association between the imputed cell proportions with measured IL-6 in both LBC1936 (n=875) and GS (n=417). In LBC1936 the multiple R^2^ statistic (representing the difference between a model adjusted for age and sex and a model adjusted for age, sex and the six cell proportions) was 0.042 and in GS it was 0.087. In GS, the multiple R^2^ for the DNAm IL-6 score was 0.53, suggesting a greater proportion of variance was explained by the cell proportions in the methylation score compared to serum IL-6.

### 3.5 Association with age in GS

As chronic inflammation is a hallmark of older age, we examined the association between the measured IL-6 phenotype and the DNAm IL-6 score with chronological age. Plots of these associations are presented in **Figure 2**. Both serum IL-6 and the DNAm IL-6 score were found to increase as a function of age (serum IL-6: β=0.022, SE=0.004 p=1.3 × 10^−7^; DNAm IL-6 score: β=0.016, SE=0.0009, p<2 × 10^−16^). No significant effect of sex was identified in the serum IL-6 dynamics; however males were found to have a higher DNAm IL-6 score compared to females (β=0.16, SE=0.02, p=3.4 × 10^−12^). Furthermore, inclusion of an interaction term between age and sex indicated the DNAm IL-6 score increased more rapidly over time in males compared to females (β=0.015, SE=0.002, p=5.1 × 10^−16^).

**Figure 2.**
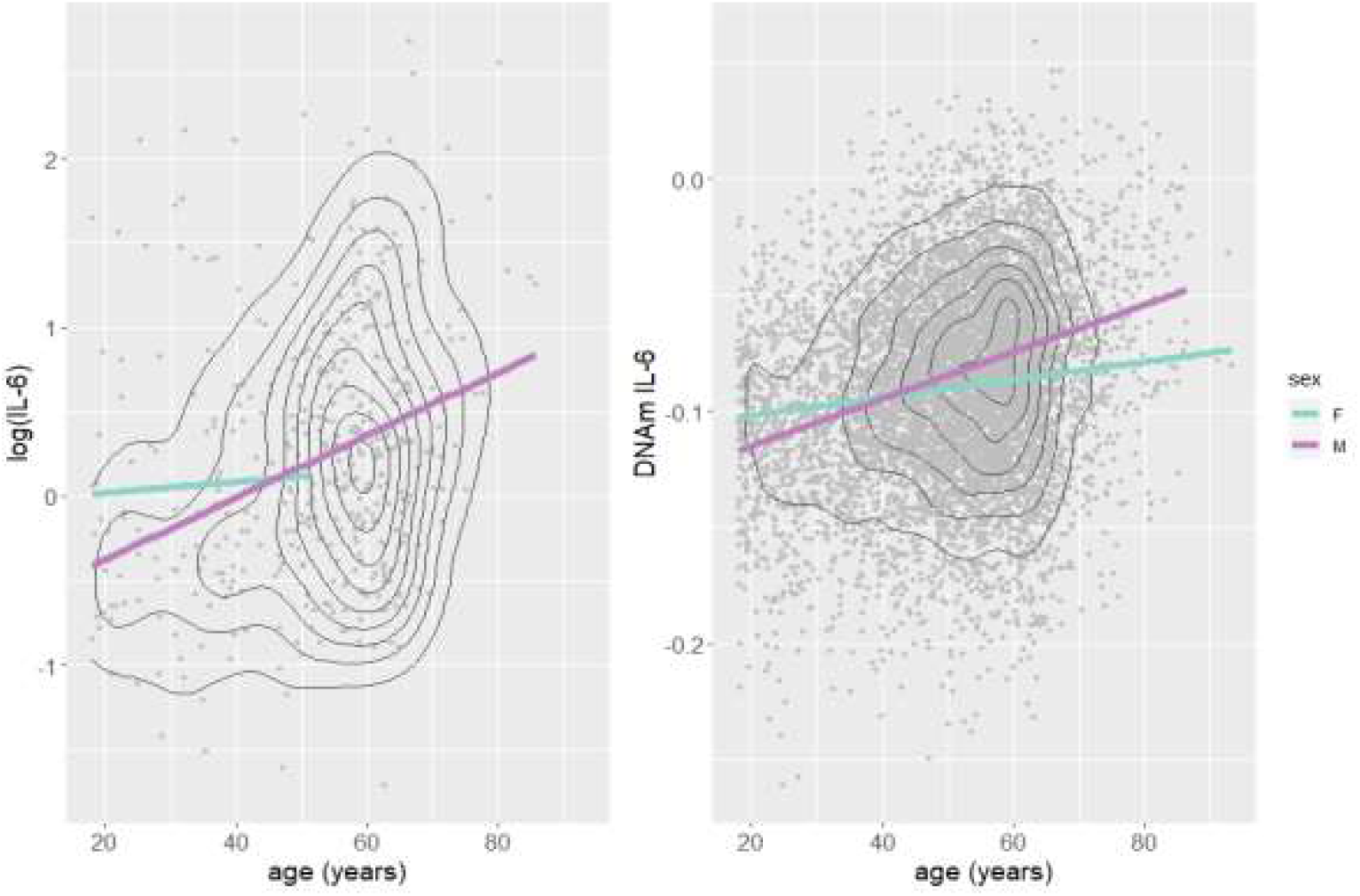
Plots of log(IL-6) (n=417) and DNAm IL-6 score (n=7,028) with chronological age in Generation Scotland. The measured IL-6 samples had previously been selected as father/offspring pairs limiting the number of samples from older females in the dataset. Grey dots represent individual participant measurements with regression lines shown for females and males in purple and blue, respectively. Black lines represent density.

### 3.6 Association with IL-6 correlates in GS

The associations between both serum log(IL-6) and the DNAm IL-6 score with factors previously associated with IL-6 levels are presented in **Table 2**.

**Table 2.**
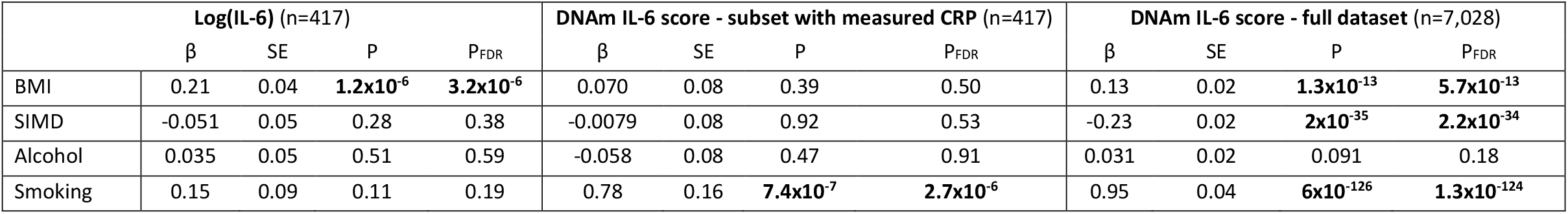
Associations between traits previously correlated with circulating IL-6, with both measured log(IL-6) and the DNAm IL-6 score in Generation Scotland. Log odds are presented for smoking. Significant associations are highlighted in bold. BMI=body mass index; SIMD=Scottish Index of Multiple Deprivation.

|  | Log(IL-6) (n=417) |  |  |  | DNAm IL-6 score - subset with measured CRP (n=417) |  |  |  | DNAm IL-6 score - full dataset (n=7,028) |  |  |  |
| --- | --- | --- | --- | --- | --- | --- | --- | --- | --- | --- | --- | --- |
| | $\beta$ | SE | P | P<sub>FDR</sub> | $\beta$ | SE | P | P<sub>FDR</sub> | $\beta$ | SE | P | P<sub>FDR</sub> |
| BMI | 0.21 | 0.04 | **1.2x10<sup>-6</sup>** | **3.2x10<sup>-6</sup>** | 0.070 | 0.08 | 0.39 | 0.50 | 0.13 | 0.02 | **1.3x10<sup>-13</sup>** | **5.7x10<sup>-13</sup>** |
| SIMD | -0.051 | 0.05 | 0.28 | 0.38 | -0.0079 | 0.08 | 0.92 | 0.53 | -0.23 | 0.02 | **2x10<sup>-35</sup>** | **2.2x10<sup>-34</sup>** |
| Alcohol | 0.035 | 0.05 | 0.51 | 0.59 | -0.058 | 0.08 | 0.47 | 0.91 | 0.031 | 0.02 | 0.091 | 0.18 |
| Smoking | 0.15 | 0.09 | 0.11 | 0.19 | 0.78 | 0.16 | **7.4x10<sup>-7</sup>** | **2.7x10<sup>-6</sup>** | 0.95 | 0.04 | **6x10<sup>-126</sup>** | **1.3x10<sup>-124</sup>** |

In the full GS cohort (n=7,028), the DNAm IL-6 score was positively associated with BMI (β=0.13, SE=0.02, p_FDR_=5.7 × 10^−13^) and self-reported smoking status (log odds=0.95, SE=0.04, p_FDR_=1.3 × 10^−124^) and negatively with social deprivation, such that a higher DNAm IL-6 score associated with a more deprived SIMD status (β=-0.23, SE=0.02, p_FDR_=2.2 × 10^−34^). No association was identified between the DNAm IL-6 score and alcohol intake (p=0.09).

In the subset of the cohort with measured IL-6 (n=417), the cytokine was positively associated with BMI (β=0.21, SE=0.04, p=3.2 × 10^−6^), but not with self-reported smoking status, social deprivation or alcohol intake (p≥0.19). In this subset, the association between the DNAm IL-6 score and smoking status remained (log odds: 0.78, SE=0.16, p_FDR_=2.7 × 10^−6^), but the associations with BMI and social deprivation were no longer significant (p≥0.5).

Given the composition of the DNAm IL-6 score, the associations with smoking status were unsurprising. As a sensitivity analysis, we re-ran the models assessing the relationship with BMI and social deprivation adjusting for methylation-predicted smoking to test for potential confounding. Here, the association with BMI was strengthened (β=0.19, SE=0.02, p_FDR_=8 × 10^−21^, increase = 46%) whereas the association with social deprivation was attenuated but remained significant (β=-0.09, SE=0.02, p_FDR_=5 × 10^−6^, attenuation = 43%).

### 3.7 Association with cognitive ability in GS

An inverse association between cognitive ability and the DNAm IL-6 score was identified in the full GS cohort (n=7,028, β=-0.14, SE=0.02, p_FDR_=1.5 × 10^−14^). Following adjustment for methylation-based smoking the association was attenuated but remained significant (β=-0.07, SE=0.02, p_FDR_=4.6 × 10^−5^, attenuation = 50%).

In the subset of the cohort with serum IL-6 data (n=417), IL-6 itself was not significantly associated with cognitive ability (β=-0.06, SE=0.05, p=0.19). Though a larger effect size was evident in the association between cognitive ability and the DNAm IL-6 score compared to IL-6 in this subset, this association was not found to be significant (n=417, β=-0.11, SE=0.07, p=0.11).

## 4. DISCUSSION

In the current study we created a poly-epigenetic score of the pro-inflammatory cytokine IL-6 based on results from an elastic net penalised regression. This DNAm IL-6 score was found to increase with age, and associate with IL-6 correlates and general fluid-type cognitive ability.

The DNAm IL-6 score highlights the potential utility of exploiting methylation data to predict biological outcomes. The magnitude of the association between the DNAm IL-6 score and serum IL-6 was modest; however, given its temporal variability, IL-6 itself may be an unreliable signature of chronic inflammation, and if the composite epigenetic score ameliorates this measure, this result would be expected. Integration of information from multiple CpG sites is potentially more likely to provide a more reliable estimate of chronic inflammation, overcoming the periodic instability of the cytokine itself. Work to investigate how the DNAm score alters in acute and chronic inflammatory conditions is warranted to assess its accuracy in this regard. We identified strong correlations between the estimated leukocyte proportions and the DNAm IL-6 score, suggesting a relatively large proportion of the variance in the latter is explained by the former. This is perhaps unsurprising given the interplay between inflammation and the immune system. IL-6 is synthesised by multiple cell types including neutrophils, B cells and macrophages in response to infectious or immunologic stimuli. Further, IL-6 enacts a pleiotropic effect on the immune response, including inducing differentiation of B cells into antibody producing cells (48), CD4^+^T cells into subsets of effector T helper cells (49), and CD8^+^T cells into cytotoxic T cells (50). However, the multiple R^2^ from the models assessing the association with measured IL-6 were small in comparison, suggesting a weaker relationship between the cytokine and the imputed immune cells. This indicates a contrast between serum IL-6 and the DNAm IL-6 score, with the methylation score more directly tracking alterations in cell proportions. Determining the causal pathway of this association is challenging given that both the DNAm IL-6 score and cell proportions were predicted from the methylation data. Assessing the relationship between the DNAm IL-6 score and measured cell counts would be beneficial to establish if the association identified here was replicated.

We identified a rise in the DNAm IL-6 score with age, with males exhibiting an accelerated trajectory compared to females. Though this sex effect was not identified in the serum IL-6 pseudo-trajectories, it has been reported previously (51, 52), and may have been missed here due to the lack of IL-6 data available from older females in the GS cohort. The DNAm IL-6 score showed associations with BMI, social deprivation and smoking status – all established correlates of IL-6. The robust association identified with smoking status was unsurprising given the inclusion of the smoking-associated probes in the DNAm IL-6 score. In the sensitivity analysis, the association with social deprivation was partly attenuated when adjusting for the methylation-based smoking score; however it remained significant and no confounding was identified in the association with BMI. No association was identified between the DNAm IL-6 score and alcohol intake which has previously been associated with IL-6 levels; however, this effect may be more acute at the time of alcohol consumption, rather than arising as a function of average units consumed (31, 53).

Though inflammation has previously been associated with cognitive ability and decline, there remains much heterogeneity in the literature assessing the relationship (54). A potential reason for this is the aforementioned acute nature of inflammatory mediators and differences in assays used to quantify them. We found that the DNAm IL-6 score associated with general cognitive ability in the full GS cohort (n=7,028). This echoes previous findings in the same cohort with a similarly-constructed DNAm score for CRP, in which no association was identified between serum CRP and cognitive ability, but an inverse association was found between the latter and the DNAm CRP score (55). This further demonstrates the potential value and applicability of proxy methylation-based scores for phenotypes that are either not available, or may in themselves be relatively unreliable. It seems the association with cognitive ability was, at least in part, driven by smoking, which has previously been associated with poorer cognitive health (56). However, though attenuated, the association did remain significant upon adjustment for the methylation-based smoking score, suggesting an independent association with the DNAm IL-6 score. Further investigation of these associations in older cohorts and those with longitudinal measures available would help to clarify the relationship, and establish if the DNAm IL-6 score associates with cognitive decline.

This is the first study to develop and characterise a DNAm-based predictor of the pro-inflammatory cytokine IL-6. The utility of this proxy phenotype is particularly evident for studies in which DNA methylation is quantified but for which IL-6 is unavailable. Additionally, utilising a composite methylation score that integrates information from multiple sites, could potentially provide a more reliable estimate of chronic inflammation, allowing for clearer insight when assessing its relation to health outcomes. This study is limited by the relatively small number of serum IL-6 samples available in the GS cohort, particularly from older females. Further, the DNAm IL-6 score was devised in a cohort with a restricted age-range all of whom have survived into the eighth decade. This may have limited the ability of the score to predict into a cohort with a broader age range and, potentially, more variable health. To further validate the DNAm IL-6 score, assessing its relationship to serum IL-6 in cohorts with a greater number of measures of the cytokine, and with repeat measures from the same individual, would be valuable.

In summary, we have illustrated that utilising DNA methylation data can provide a proxy for the pro-inflammatory cytokine IL-6 that associates with pertinent health, lifestyle, and cognitive, outcomes. Future studies with DNA methylation data, inflammatory disease measures, and longitudinal follow-up available will allow for further insight into the utility of this DNAm-based predictor and its ability to act as a measure of chronic inflammation compared to the labile phenotype.

## 5. Data availability

LBC data are available on request from the Lothian Birth Cohort Study, University of Edinburgh (Ian Deary). LBC data are not publicly available due to them containing information that could compromise participant consent and confidentiality. According to the terms of consent for Generation Scotland participants, access to data must be reviewed by the Generation Scotland Access Committee. Applications should be made to.

## 6. Acknowledgements

The authors thank all LBC1936 study participants and research team members who have contributed, and continue to contribute, to ongoing studies. LBC1936 is supported by Age UK (Disconnected Mind program) and the Medical Research Council (MR/M01311/1). Methylation typing was supported by Centre for Cognitive Ageing and Cognitive Epidemiology (Pilot Fund award), Age UK, The Wellcome Trust Institutional Strategic Support Fund, The University of Edinburgh, and The University of Queensland. The Olink^®^ Proteomics assays were supported by a National Institutes of Health (NIH) research grant R01AG054628. This work was in part conducted in the Centre for Cognitive Ageing and Cognitive Epidemiology, which is supported by the Medical Research Council and Biotechnology and Biological Sciences Research Council (MR/K026992/1) and which supports IJD.

The authors thank all individuals and project team members who have contributed to both GS and to the ‘STRADL: Stratifying Resilience and Depression Longitudinally’ follow-up study. GS received core support from the Chief Scientist Office of the Scottish Government Health Directorates (CZD/16/6) and the Scottish Funding Council (HR03006) and is currently supported by the Wellcome Trust (216767/Z/19/Z). Genotyping and DNA methylation profiling of the GS samples was carried out by the Genetics Core Laboratory at the Wellcome Trust Clinical Research Facility, Edinburgh, Scotland and was funded by the Medical Research Council UK and the Wellcome Trust (Wellcome Trust Strategic Award “STratifying Resilience and Depression Longitudinally” ((STRADL) Reference 104036/Z/14/Z)).

AJS, DAG and RFH are Translational Neuroscience PhD students funded by Wellcome (203771/Z/16/Z to AJS; 108890/Z/15/Z to DAG and RFH). REM and DLM_C_C are supported by an Alzheimer’s Research UK major project grant (ARUK-PG2017B-10). TSJ is supported by the European Research Council (ERC) under the European Union’s Horizon 2020 research and innovation programme (Grant agreement No. 681181) and the UK Dementia Research Institute which receives its funding from DRI Ltd, funded by the UK Medical Research Council, Alzheimer’s Society, and Alzheimer’s Research UK. PMV is supported by the Australian National Health and Medical Research Council (1113400) and the Australian Research Council (FL180100072).

---

**Supplementary Table 1.** Elastic net regression CpGs and corresponding weights for IL-6

| *cpg* | *coefficient* |
| --- | --- |
| cg05575921 | -0.056521923 |
| cg03885055 | -0.160772495 |
| cg17412005 | -0.320303401 |
| cg04921989 | 0.053893633 |
| cg14965639 | -0.107877072 |
| cg20789595 | -0.192768423 |
| cg19584649 | 0.121457246 |
| cg25250132 | 0.211296897 |
| cg23729763 | 0.14808051 |
| cg20059928 | -0.014278955 |
| cg04928129 | -0.613477882 |
| cg12929678 | 0.171948359 |

**Supplementary Figure 1.**
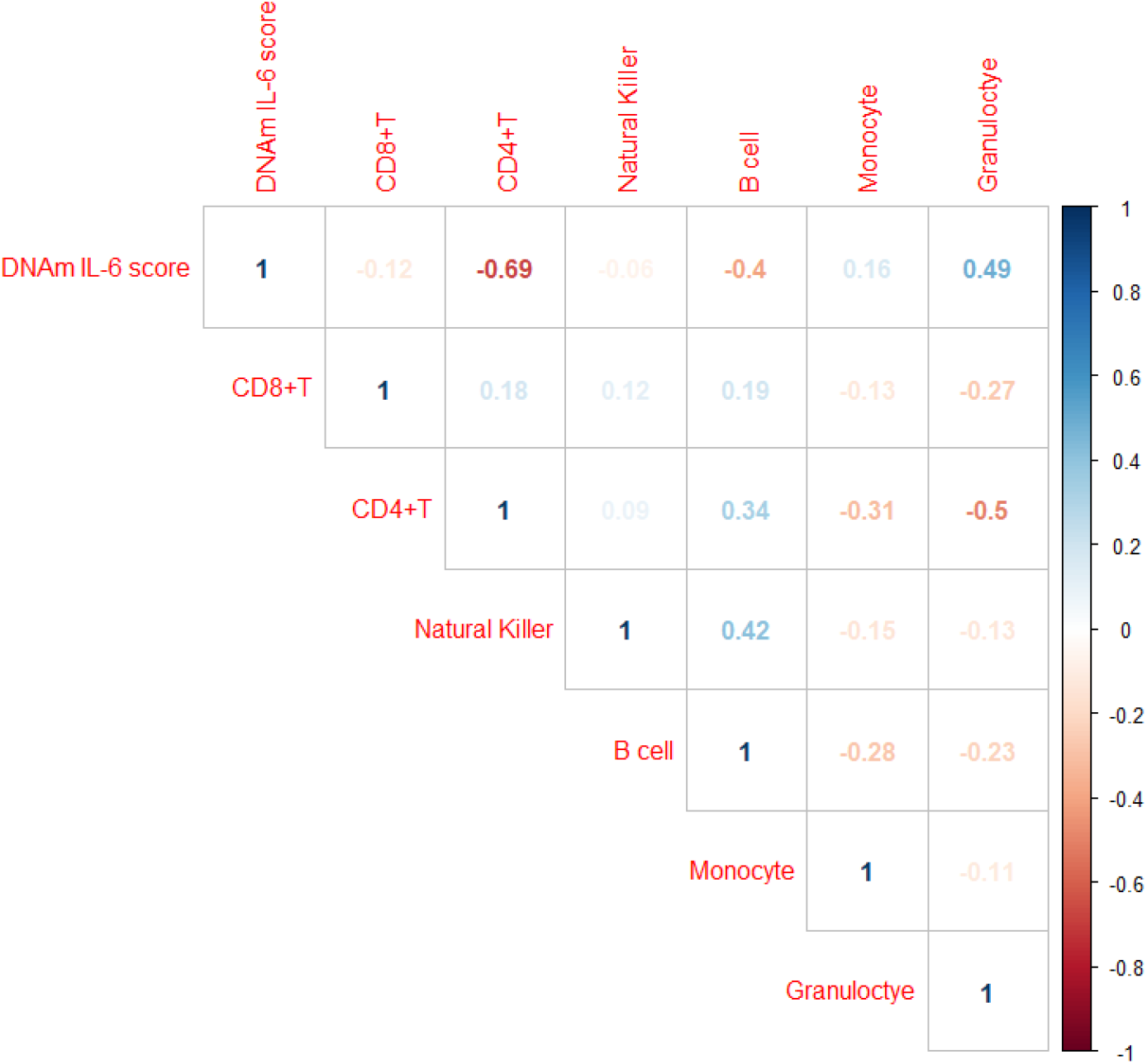
Pearson correlations between the DNAm IL-6 score and the imputed cell proportions in Generation Scotland.

**Supplementary Figure 2.**
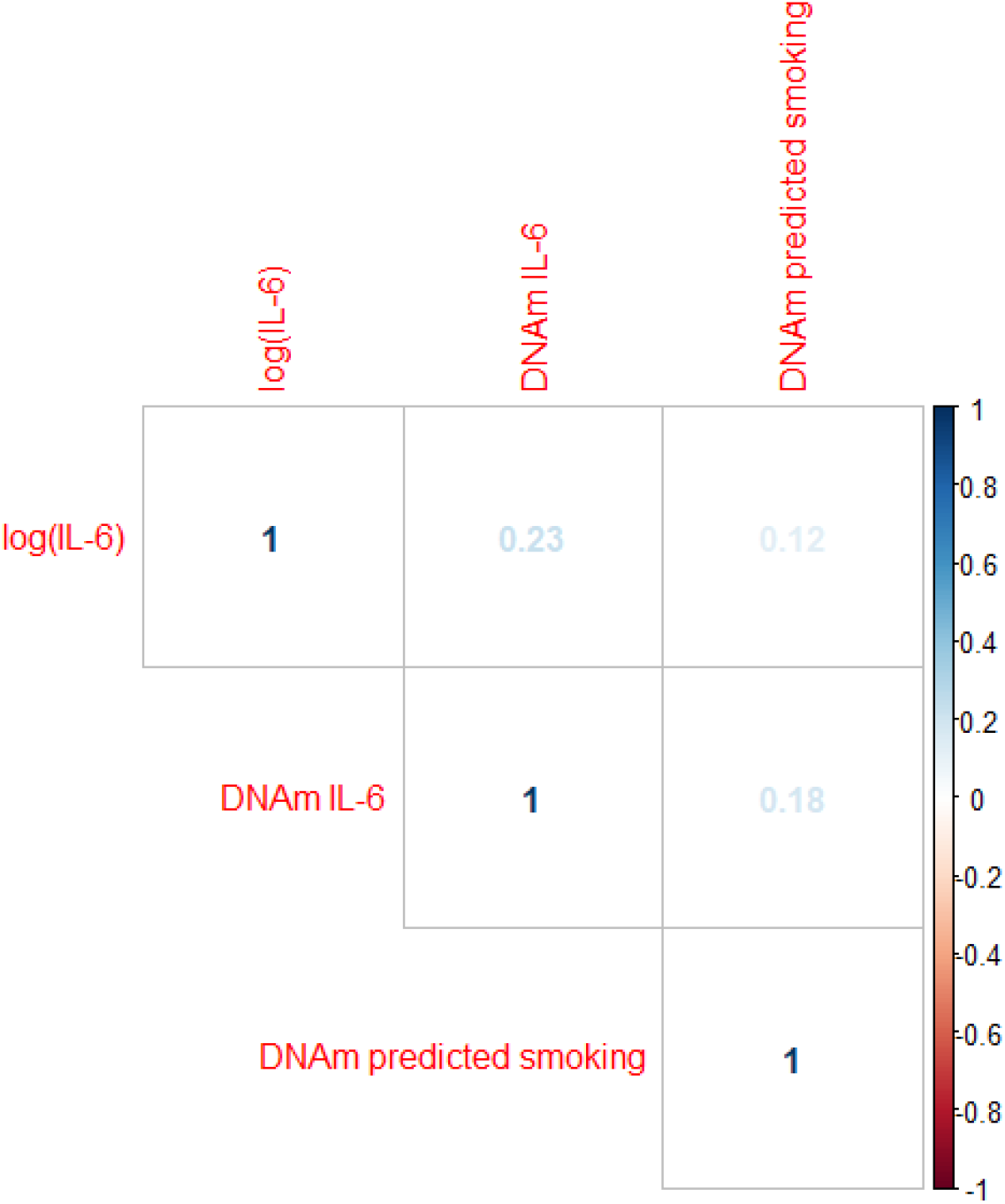
Pearson correlations between measured log(IL-6), the DNAm IL-6 score and the DNA methylation-based smoking score in Generation Scotland.

## REFERENCES

1. Kinney JW, Bemiller SM, Murtishaw AS, Leisgang AM, Salazar AM, Lamb BT. Inflammation as a central mechanism in Alzhei. Alzheimers Dement (N Y). 2018;4:575–90.

2. Greten FR, Grivennikov SI. Inflammation and Cancer: Triggers, Mechanisms, and Consequences. Immunity. 2019;51(1):27–41.

3. Chung HY, Cesari M, Anton S, Marzetti E, Giovannini S, Seo AY, Carter C, Yu BP, Leeuwenburgh C. Molecular inflammation: underpinnings of aging and age-related diseases. Ageing research reviews. 2009;8(1):18–30.

4. Conti P, Shaik-Dasthagirisaeb Y. Atherosclerosis: a chronic inflammatory disease mediated by mast cells. Cent Eur J Immunol. 2015;40(3):380–6.

5. Results of a follow-up study to the randomized Alzheimer’s Disease Anti-inflammatory Prevention Trial (ADAPT). Alzheimer’s & dementia : the journal of the Alzheimer’s Association. 2013;9(6):714–23.

6. Arvanitakis Z, Grodstein F, Bienias JL, Schneider JA, Wilson RS, Kelly JF, Evans DA, Bennett DA. Relation of NSAIDs to incident AD, change in cognitive function, and AD pathology. Neurology. 2008;70(23):2219–25.

7. Heinrich PC, Castell JV, Andus T. Interleukin-6 and the acute phase response. The Biochemical journal. 1990;265(3):621–36.

8. Gabay C. Interleukin-6 and chronic inflammation. Arthritis Res Ther. 2006;8 Suppl 2(Suppl 2):S3–S.

9. Moldoveanu AI, Shephard RJ, Shek PN. Exercise elevates plasma levels but not gene expression of IL-1β, IL-6, and TNF-α in blood mononuclear cells. urnal of Applied Physiology. 2000;89(4):1499–504.

10. Lundman P, Boquist S, Samnegård A, Bennermo M, Held C, Ericsson CG, Silveira A, Hamsten A, Tornvall P. A high-fat meal is accompanied by increased plasma interleukin-6 concentrations. Nutrition, Metabolism and Cardiovascular Diseases. 2007;17(3):195–202.

11. Dugué B, Leppänen E. Short-term variability in the concentration of serum interleukin-6 and its soluble receptor in subjectively healthy persons. Clinical Chemistry and Laboratory Medicine. 1998;36(5):323–5.

12. Cava F, González C, Pascual MJ, Navajo JA, González-Buitrago Jm. BIOLOGICAL VARIATION OF INTERLEUKIN 6 (IL-6) AND SOLUBLE INTERLEUKIN 2 RECEPTOR (sIL2R) IN SERUM OF HEALTHY INDIVIDUALS. Cytokine. 2000;12(9):1423–5.

13. Jones PA. Functions of DNA methylation: islands, start sites, gene bodies and beyond. Nature reviews Genetics. 2012;13(7):484–92.

14. Stenvinkel P, Karimi M, Johansson S, Axelsson J, Suliman M, Lindholm B, Heimburger O, Barany P, Alvestrand A, Nordfors L, Qureshi AR, Ekstrom TJ, Schalling M. Impact of inflammation on epigenetic DNA methylation - a novel risk factor for cardiovascular disease? Journal of internal medicine. 2007;261(5):488–99.

15. Hartnett L, Egan LJ. Inflammation, DNA methylation and colitis-associated cancer. Carcinogenesis. 2012;33(4):723–31.

16. Nilsson E, Jansson PA, Perfilyev A, Volkov P, Pedersen M, Svensson MK, Poulsen P, Ribel-Madsen R, Pedersen NL, Almgren P, Fadista J, Ronn T, Klarlund Pedersen B, Scheele C, Vaag A, Ling C. Altered DNA methylation and differential expression of genes influencing metabolism and inflammation in adipose tissue from subjects with type 2 diabetes. Diabetes. 2014;63(9):2962–76.

17. Ligthart S, Marzi C, Aslibekyan S, Mendelson MM, Conneely KN, Tanaka T, Colicino E, Waite LL, Joehanes R, Guan W, Brody JA, Elks C, Marioni R, Jhun MA, Agha G, Bressler J, Ward-Caviness CK, Chen BH, Huan T, Bakulski K, Salfati EL, Fiorito G, Wahl S, Schramm K, Sha J, Hernandez DG, Just AC, Smith JA, Sotoodehnia N, Pilling LC, Pankow JS, Tsao PS, Liu C, Zhao W, Guarrera S, Michopoulos VJ, Smith AK, Peters MJ, Melzer D, Vokonas P, Fornage M, Prokisch H, Bis JC, Chu AY, Herder C, Grallert H, Yao C, Shah S, McRae AF, Lin H, Horvath S, Fallin D, Hofman A, Wareham NJ, Wiggins KL, Feinberg AP, Starr JM, Visscher PM, Murabito JM, Kardia SLR, Absher DM, Binder EB, Singleton AB, Bandinelli S, Peters A, Waldenberger M, Matullo G, Schwartz JD, Demerath EW, Uitterlinden AG, van Meurs Jbj, Franco OH, Chen Y-DI, Levy D, Turner ST, Deary IJ, Ressler KJ, Dupuis J, Ferrucci L, Ong KK, Assimes TL, Boerwinkle E, Koenig W, Arnett DK, Baccarelli AA, Benjamin EJ, Dehghan A, Investigators W-E, Disease CeoCH. DNA methylation signatures of chronic low-grade inflammation are associated with complex diseases. Genome Biology. 2016;17(1):255.

18. Myte R, Sundkvist A, Van Guelpen B, Harlid S. Circulating levels of inflammatory markers and DNA methylation, an analysis of repeated samples from a population based cohort. Epigenetics. 2019;14(7):649–59.

19. Verschoor CP, McEwen LM, Kohli V, Wolfson C, Bowdish DM, Raina P, Kobor MS, Balion C. The relation between DNA methylation patterns and serum cytokine levels in community-dwelling adults: a preliminary study. BMC Genet. 2017;18(1):57-.

20. McCartney DL, Hillary RF, Stevenson AJ, Ritchie SJ, Walker RM, Zhang Q, Morris SW, Bermingham ML, Campbell A, Murray AD, Whalley HC, Gale CR, Porteous DJ, Haley CS, McRae AF, Wray NR, Visscher PM, McIntosh AM, Evans KL, Deary IJ, Marioni RE. Epigenetic prediction of complex traits and death. Genome Biology. 2018;19(1):136.

21. Deary IJ, Gow AJ, Taylor MD, Corley J, Brett C, Wilson V, Campbell H, Whalley LJ, Visscher PM, Porteous DJ, Starr JM. The Lothian Birth Cohort 1936: a study to examine influences on cognitive ageing from age 11 to age 70 and beyond. BMC geriatrics. 2007;7:28.

22. Taylor AM, Pattie A, Deary IJ. Cohort Profile Update: The Lothian Birth Cohorts of 1921 and 1936. International journal of epidemiology. 2018;47(4):1042–r.

23. Zhang Q, Marioni RE, Robinson MR, Higham J, Sproul D, Wray NR, Deary IJ, McRae AF, Visscher PM. Genotype effects contribute to variation in longitudinal methylome patterns in older people. Genome medicine. 2018;10(1):75.

24. Shah S, McRae AF, Marioni RE, Harris SE, Gibson J, Henders AK, Redmond P, Cox SR, Pattie A, Corley J, Murphy L, Martin NG, Montgomery GW, Starr JM, Wray NR, Deary IJ, Visscher PM. Genetic and environmental exposures constrain epigenetic drift over the human life course. Genome Res. 2014;24(11):1725–33.

25. Friedman JH, Hastie T, Tibshirani R. Regularization Paths for Generalized Linear Models via Coordinate Descent. Journal of Statistical Software; Vol 1, Issue 1 (2010). 2010.

26. Smith BH, Campbell A, Linksted P, Fitzpatrick B, Jackson C, Kerr SM, Deary IJ, Macintyre DJ, Campbell H, McGilchrist M, Hocking LJ, Wisely L, Ford I, Lindsay RS, Morton R, Palmer CN, Dominiczak AF, Porteous DJ, Morris AD. Cohort Profile: Generation Scotland: Scottish Family Health Study (GS:SFHS). The study, its participants and their potential for genetic research on health and illness. International journal of epidemiology. 2013;42(3):689–700.

27. McCartney DL, Stevenson AJ, Walker RM, Gibson J, Morris SW, Campbell A, Murray AD, Whalley HC, Porteous DJ, McIntosh AM, Evans KL, Deary IJ, Marioni RE. Investigating the relationship between DNA methylation age acceleration and risk factors for Alzheimer’s disease. Alzheimers Dement (Amst). 2018;10:429–37.

28. Madden RA, McCartney DL, Walker RM, Hillary RF, Bermingham ML, Rawlik K, Morris SW, Campbell A, Porteous DJ, Deary IJ, Evans KL, Hafferty J, McIntosh AM, Marioni RE. Birth weight predicts psychiatric and physical health, cognitive function, and DNA methylation differences in an adult population. bioRxiv. 2019:664045.

29. Sunyer J, Forastiere F, Pekkanen J, Plana E, Kolz M, Pistelli R, Jacquemin B, Brüske-Hohlfeld I, Pitsavos C, Bellander T, Koenig W, Peters A. Interaction between smoking and the interleukin-6 gene affects systemic levels of inflammatory biomarkers. Nicotine & tobacco research : official journal of the Society for Research on Nicotine and Tobacco. 2009;11(11):1347–53.

30. Helmersson J, Larsson A, Vessby B, Basu S. Active smoking and a history of smoking are associated with enhanced prostaglandin F2α, interleukin-6 and F2-isoprostane formation in elderly men. Atherosclerosis. 2005;181(1):201–7.

31. González-Quintela A, Dominguez-Santalla MJ, Pérez LF, Vidal C, Lojo S, Barrio E. INFLUENCE OF ACUTE ALCOHOL INTAKE AND ALCOHOL WITHDRAWAL ON CIRCULATING LEVELS OF IL-6, IL-8, IL-10 AND IL-12. Cytokine. 2000;12(9):1437–40.

32. Sindhu S, Thomas R, Shihab P, Sriraman D, Behbehani K, Ahmad R. Obesity Is a Positive Modulator of IL-6R and IL-6 Expression in the Subcutaneous Adipose Tissue: Significance for Metabolic Inflammation. PLoS One. 2015;10(7):e0133494–e.

33. Khaodhiar L, Ling PR, Blackburn GL, Bistrian BR. Serum levels of interleukin-6 and C-reactive protein correlate with body mass index across the broad range of obesity. JPEN Journal of parenteral and enteral nutrition. 2004;28(6):410–5.

34. Petersen KL, Marsland AL, Flory J, Votruba-Drzal E, Muldoon MF, Manuck SB. Community socioeconomic status is associated with circulating interleukin-6 and C-reactive protein. Psychosomatic medicine. 2008;70(6):646–52.

35. de Britto Rosa NM, de Queiroz BZ, Pereira DS, di Sabatino Santos Mla, Oliveira DMG, Narciso FMeS, Pereira LSM. Interleukin-6 plasma levels and socioeconomic status in Brazilian elderly community-dwelling women. Archives of Gerontology and Geriatrics. 2011;53(2):196–9.

36. Bollepalli S, Korhonen T, Kaprio J, Anders S, Ollikainen M. EpiSmokEr: a robust classifier to determine smoking status from DNA methylation data. Epigenomics. 2019;11(13):1469–86.

37. Elliott HR, Tillin T, McArdle WL, Ho K, Duggirala A, Frayling TM, Davey Smith G, Hughes AD, Chaturvedi N, Relton CL. Differences in smoking associated DNA methylation patterns in South Asians and Europeans. Clinical Epigenetics. 2014;6(1):4.

38. Zeilinger S, Kühnel B, Klopp N, Baurecht H, Kleinschmidt A, Gieger C, Weidinger S, Lattka E, Adamski J, Peters A, Strauch K, Waldenberger M, Illig T. Tobacco Smoking Leads to Extensive Genome-Wide Changes in DNA Methylation. PLoS One. 2013;8(5):e63812.

39. Marioni RE, Campbell A, Scotland G, Hayward C, Porteous DJ, Deary IJ. Differential effects of the APOE e4 allele on different domains of cognitive ability across the life-course. European journal of human genetics : EJHG. 2016;24(6):919–23.

40. Hagenaars SP, Harris SE, Clarke T-K, Hall L, Luciano M, Fernandez-Pujals AM, Davies G, Hayward C, Generation S, Starr JM, Porteous DJ, McIntosh AM, Deary IJ. Polygenic risk for coronary artery disease is associated with cognitive ability in older adults. International journal of epidemiology. 2016;45(2):433–40.

41. R Core Team. R: A Language and Environment for Statisticl Computing. R Foundation for Statistical Computng, Vienna, Austria. URL https://www.R-project.org/. 2019.

42. Philibert RA, Beach SR, Brody GH. Demethylation of the aryl hydrocarbon receptor repressor as a biomarker for nascent smokers. Epigenetics. 2012;7(11):1331–8.

43. Fasanelli F, Baglietto L, Ponzi E, Guida F, Campanella G, Johansson M, Grankvist K, Johansson M, Assumma MB, Naccarati A. Hypomethylation of smoking-related genes is associated with future lung cancer in four prospective cohorts. Nature communications. 2015;6(1):1–9.

44. Zhu X, Li J, Deng S, Yu K, Liu X, Deng Q, Sun H, Zhang X, He M, Guo H. Genome-wide analysis of DNA methylation and cigarette smoking in a Chinese population. Environmental health perspectives. 2016;124(7):966–73.

45. Harlid S, Xu Z, Panduri V, Sandler DP, Taylor JA. CpG sites associated with cigarette smoking: analysis of epigenome-wide data from the sister study. Environmental health perspectives. 2014;122(7):673–8.

46. Joehanes R, Just AC, Marioni RE, Pilling LC, Reynolds LM, Mandaviya PR, Guan W, Xu T, Elks CE, Aslibekyan S, Moreno-Macias H, Smith JA, Brody JA, Dhingra R, Yousefi P, Pankow JS, Kunze S, Shah SH, McRae AF, Lohman K, Sha J, Absher DM, Ferrucci L, Zhao W, Demerath EW, Bressler J, Grove ML, Huan T, Liu C, Mendelson MM, Yao C, Kiel DP, Peters A, Wang-Sattler R, Visscher PM, Wray NR, Starr JM, Ding J, Rodriguez CJ, Wareham NJ, Irvin MR, Zhi D, Barrdahl M, Vineis P, Ambatipudi S, Uitterlinden AG, Hofman A, Schwartz J, Colicino E, Hou L, Vokonas PS, Hernandez DG, Singleton AB, Bandinelli S, Turner ST, Ware EB, Smith AK, Klengel T, Binder EB, Psaty BM, Taylor KD, Gharib SA, Swenson BR, Liang L, DeMeo DL, O’Connor GT, Herceg Z, Ressler KJ, Conneely KN, Sotoodehnia N, Kardia SLR, Melzer D, Baccarelli AA, van Meurs Jbj, Romieu I, Arnett DK, Ong KK, Liu Y, Waldenberger M, Deary IJ, Fornage M, Levy D, London SJ. Epigenetic Signatures of Cigarette Smoking. Circ Cardiovasc Genet. 2016;9(5):436–47.

47. Chatziioannou A, Georgiadis P, Hebels DG, Liampa I, Valavanis I, Bergdahl IA, Johansson A, Palli D, Chadeau-Hyam M, Siskos AP, Keun H, Botsivali M, de Kok TM, Pérez AE, Kleinjans JC, Vineis P, Kyrtopoulos SA. Blood-based omic profiling supports female susceptibility to tobacco smoke-induced cardiovascular diseases. Scientific reports. 2017;7:42870.

48. Kishimoto T. Factors affecting B-cell growth and differentiation. Annu Rev Immunol. 1985;3:133–57.

49. Dienz O, Rincon M. The effects of IL-6 on CD4 T cell responses. Clin Immunol. 2009;130(1):27–33.

50. Okada M, Kitahara M, Kishimoto S, Matsuda T, Hirano T, Kishimoto T. IL-6/BSF-2 functions as a killer helper factor in the in vitro induction of cytotoxic T cells. J Immunol. 1988;141(5):1543–9.

51. Milan-Mattos JC, Anibal FF, Perseguini NM, Minatel V, Rehder-Santos P, Castro CA, Vasilceac FA, Mattiello SM, Faccioli LH, Catai AM. Effects of natural aging and gender on pro-inflammatory markers. Braz J Med Biol Res. 2019;52(9):e8392–e.

52. Zhu S, Patel KV, Bandinelli S, Ferrucci L, Guralnik JM. Predictors of interleukin-6 elevation in older adults. J Am Geriatr Soc. 2009;57(9):1672–7.

53. Neupane SP, Skulberg A, Skulberg KR, Aass HCD, Bramness JG. Cytokine Changes following Acute Ethanol Intoxication in Healthy Men: A Crossover Study. Mediators of Inflammation. 2016;2016:3758590.

54. Bettcher BM, Kramer JH. Longitudinal inflammation, cognitive decline, and Alzheimer’s disease: a mini-review. Clin Pharmacol Ther. 2014;96(4):464–9.

55. Stevenson AJ, McCartney DL, Hillary RF, Campbell A, Morris SW, Bermingham ML, Walker RM, Evans KL, Boutin TS, Hayward C, McRae AF, McColl BW, Spires-Jones TL, McIntosh AM, Deary IJ, Marioni RE. Characterisation of an inflammation-related epigenetic score and its association with cognitive ability. bioRxiv. 2019:802009.

56. Nooyens AC, van Gelder BM, Verschuren WM. Smoking and cognitive decline among middle-aged men and women: the Doetinchem Cohort Study. American journal of public health. 2008;98(12):2244–50.

